# Narratives of the convalescent plasma donor in a Peruvian social security hospital.: motivations, fears, expectations and experiences

**DOI:** 10.1101/2022.02.16.22270690

**Authors:** Silvana M. Matassini Eyzaguirre, Christian Villanueva Yapa, Ausberto Chunga Chunga, Arturo Sagastegui Soto, Ibeth Melania Neyra Vera, Suly Soto Ordoñez, Martina Guillermo Román, Martin Oyanguren Miranda, Percy Soto-Becerra, Leda Yamileé Hurtado Roca, Jorge L. Maguiña, Araujo-Castillo Roger Vladimir

**Author notes:** **Corresponding author:** Silvana M. Matassini Eyzaguirre. **Contributions:** All authors reviewed and approved the final version of the manuscript. **Funding:** This study was funded by the Instituto de Evaluacion de Tecnologias en Salud e Investigacion - IETSI, EsSalud, Peru. Likewise, this is an ancillary study to a clinical trial partially funded by the National Council of Science, Technology and Technological Innovation 471 (CONCYTEC) through the National Fund for Scientific, Technological Development and 472 Technological Innovation (FONDECYT), subvention (Nº068-2020-FONDECYT) by Legislative Decree Nº 1473.

## Abstract

**Objectives:** To know and explore from convalescent plasma donator’s voices the experience in the blood donation process at a Peruvian social security hospital.

**Methods:** Qualitative study with a phenomenological design. The investigation was carried out in 01 hospitals of the social security of Peru. Semi-structured interviews were carried out.

**Results:** Eleven donors of convalescent plasma were interviewed. The main motivations for donating were being able to contribute to national research and supporting patients affected by COVID-19. Fears focus on the possible risk of contagion within the hospital. Donors emphasised the attention and support of health personnel alongside the donation procedure. The main expectations and suggestions point towards greater dissemination of donation campaigns with special emphasis on safety. Likewise, an improvement in the time of the donation procedure (from enrolment to the extraction of convalescent plasma), and the implementation of friendly spaces to encourage future blood donation campaigns were highlighted.

**Conclusions:** The experience of the convalescent plasma donors was positive. However, improvements must be made in terms of processes and infrastructure to ensure future successful blood donation campaigns.

## Background

SARS-CoV-2 is the etiologic agent of coronavirus disease 2019 (COVID-19), identified in China in December 2019 [1]. The disease spread rapidly worldwide, such that WHO declared it a pandemic on March 12, 2020. Despite harsh containment measures, the spread of this virus continued worldwide. There are currently no specific treatments against COVID-19, so in the absence of a known effective therapy, the development of new therapies to treat symptomatic patients and reduce the risk of adverse outcomes became a priority.

Convalescent plasma (CP) from patients recovered from COVID-19 was one of the treatments that were considered potentially effective early in the pandemic, and several clinical trials were quickly initiated worldwide [2].Theoretically, plasma from convalescents of patients recovered from the disease contains antibodies that would help patients with COVID-19 to cure the disease or reduce its severity and mortality. However, currently, some clinical trials and meta-analyses [3,4] conclude that, in general, PC would not reduce mortality in patients with moderate disease and would have little or no effect on measures of clinical improvement. For that reason, some clinical practice guidelines and living systematic reviews do not recommend using PC for COVID-19 treatment. Nevertheless, although there is no consensus yet, PC is being used in many countries to treat COVID-19 [5].

On the other hand, some recent studies reveal that a specific type of PC, hyperimmune plasma, as opposed to ‘conventional’ PC, may indeed be an alternative treatment for COVID-19, especially if its administration occurs early [6–9]. In addition, clinical trials have found that it reduces mortality and disease progression in older adult patients and in patients who did not receive mechanical ventilation [6–9]. These promising results, still under evaluation, would indicate that hyperimmune PC could be an alternative treatment used in contexts where additional alternatives are needed.

In 2020, Peru’s social health security, EsSalud, initiated a randomized, open-label, controlled clinical trial that sought to evaluate the efficacy of PC in the treatment of patients with COVID-19 [10]. EsSalud is part of Peru’s decentralized health care system and covers 33% of the Peruvian population, especially the economically active population working in the public or formal private sector, retired pensioners, and their immediate family members [11]. In the absence of effective treatments, EsSalud expressed its intention to evaluate the use of PC to assess the possibility of implementing it according to national and international evidence. For this reason, if hyperimmune PC is shown to be effective for the treatment of COVID-19, one of the challenges faced by the Peruvian social health security is to guarantee the continued supply of PC from voluntary donors, former COVID-19 convalescent patients, in order to meet the demand for it. During the health crisis, the health system’s challenges in developing countries are more visible. The precariousness of the health system led to a collapse of services, overcrowding, an increase of the already existing bureaucratic and infrastructure barriers, etc. On the other hand, the fears, insecurities, and uncertainty of the population could hinder the willingness of the population and, therefore, the campaigns not only of convalescent plasma donation but also of blood donation in general [12]. In order to achieve greater citizen participation in this type of campaign and the face of health crises such as the current ones, it is necessary to explore the experiences of donors at the individual level and about the health system so that, from their voices, the main strengths, and weaknesses of the health system can be visualised and discussed. Furthermore, a positive and satisfactory experience of the donation will contribute to an increase in return, dissemination, and awareness of the importance of donation.

For this reason, in the context of a clinical trial, the present study aimed to explore the diverse experiences of convalescent plasma donors, which also implies evaluating the role of the donor, of the population, in situations of a global health crisis. Exploring their experiences, their expectations, their confrontations with the health system in a Peruvian social security hospital, will give us light to generate recommendations and strategies that bring the population closer to the health system and promote donation campaigns valuing, respecting and knowing the motivations, needs, and expectations of donors.

Finally, exploring the narratives of the convalescent plasma donor brings to the forefront those people who faced the disease and the health system. Therefore, now as convalescent plasma donors and with that lived experience, these people can express and share motivations, fears, needs, and expectations that dialogue with the materiality and infrastructure of social health security. Furthermore, collecting their narratives will allow decision-makers to design various intervention strategies and reinforce and improve hospital services.

## Method

This research is a sub-study of the clinical trial registered at: https://ictrptest.azurewebsites.net/Trial2.aspx?TrialID=PER-013-20.

A qualitative study of phenomenological design was conducted exploring the individual experience of convalescent plasma donors.

### Recruitment

Participants attended a social health security hospital in Lima, Peru. Recruitment of participants for the present study was done through the official EsSalud Facebook page. An invitation was disseminated to those who had participated in the process of donating convalescent plasma from SARS-CoV-2 infection, recruiting eleven people who participated in the convalescent plasma clinical trial as donors. All participants went on to complete the donation process.

Semi-structured interviews were conducted via telephone calls between February and March 2021. Before starting the data collection, a pilot test was carried out, which consisted of interviewing five people to validate the methodological tool and to define the interaction strategies with the participants in terms of relationship and time.

Participants were asked to confirm their participation by reading and signing the virtual signature (recorded verbally).

### Interviews

The interviews were conducted individually, via telephone by a specialist in social sciences, showing sensitivity to the subject matter and respecting the customs and beliefs of the population. Each interview lasted approximately 45 minutes and was recorded with the verbal consent of the participants.

The thematic axes are summarised in the next table:

**Table 1.** Thematic axes.

| **Topics** |  |
| --- | --- |
| Education | • Level of education attained |
| Health | • History of diseases (antecedents) and their confrontations.<br>• Main health problems.<br>• Frequent medical visits/consultations. |
| COVID-19-Plasma donation | • Knowledge about blood donation |
| Motivations for donating | • Individual, collective.<br>• Acknowledgments. |
| Fears | • Personal insecurities (fear about the |
|  | process on their health, exposure as a patient, etc.).<br>• Insecurities related to the health system and blood donation procedures. |
| Experiences | • Processes and perceptions from registration to blood collection. |
| Expectations | • What they expect from the health system and blood donation procedures.<br>• What they expect about the achievements and impact on the community from their participation in blood donation campaigns. |

The study was approved by the specific research ethics committee for COVID-19 of the Institute for Health Technology Assessment and Research of EsSalud (IETSI - EsSalud), on December 19, 2020.

### Analysis

The information reduction process was initiated by transcribing interviews and their subsequent coding according to the proposed thematic axes. The process of organizing and coding the information was carried out in Atlas ti version 9.0. Participants were not invited to contrast the findings of the study.

The analysis findings are accompanied by textual quotations from the participants, which will be presented with the initials of fictitious names, respecting the criteria of confidentiality and protection of the research participants. These quotations were selected due to their representativeness and similarity with the majority of the results.

## Results

All the people who wished to participate after the dissemination of the study were interviewed, nine men and two women. The ages of the participants ranged from 23-54 years. Of the total number of participants, one man and one woman were the only ones who had previously donated blood in Perú. All eleven participants had higher technical or university education. The donors were permanent residents of the city of Lima.

### Health

Of the total number of participants, 10 did not make frequent visits to hospitals or private practices and reported that they were in good health and that COVID-19 was the most worrisome illness experience they had had to date. One man in his 40s was the only one who had had to deal with life-threatening diseases (tuberculosis and hepatitis A) in previous years. According to him, these diseases did not leave any sequelae in his body.

> *“*…*the last medical consultation I had was when I was a teenager, I was still in Chiclayo and it was for acne. But really you could say that it’s been more than eight years since I’ve been to a doctor’s office”* (JM, male).
>
> *“I don’t have any history of any disease, I’ve never had any disease, I’ve never been hospitalised, I have no history. No risk conditions, either. Lately, the only history I would have now, would be that I got infected with COVID, but not after that”* (ST, female).

### COVID-19 and plasma donation

Convalescent plasma donors were optimistic about plasma donation and about blood donation in general. They highlight its potential benefits both for COVID-19 cases and for other types of diseases.

> “*Blood donation, serves to save lives and depends on what is needed, red blood cells, platelets, plasma itself, whatever is needed according to the patient’s condition”* (MR, male).
>
> *“I know that in Peru they have been donating plasma to infuse antibodies to critical patients who have been infected with COVID. I am a frequent donor of platelets and blood, so blood donation is something very important because it can help many people, it is a way to save and help people”* (ST, female).
>
> *“I think we should all help others. One thing is blood and another thing is plasma, I heard that plasma was going to cure the people they were going to treat. That’s why I say, I went through that disease and how nice that I can help people who can recover or treat COVID”* (DA, male).

### Motivations for donating

The two main reasons that drove the intention to donate convalescent plasma were (i) support for research and (ii) support for victims of COVID-19.

> *“Since here in Perú the support in research is not very good, so I said if we can support a little with research in Perú on Coronavirus, great, if you can support with plasma, great, that was one of the reasons why I also accepted”* (VR, female).
>
> *“To help other people. And if with my blood I can help more people, well, let it be done. Not for an economic incentive, or anything, if we can help other people that way, then we have to help them that way”* (JK, male).
>
> *“My motivation has been mainly to support with the issue of Coronavirus research, especially for the issue of trials. Because I understand the important role of volunteers”* (GQ, male).

Regarding recognition for their participation as donors, the participants mentioned that they were aware that they would not receive monetary compensation but that, in addition, their motivations were more related to collaboration than to any other type of incentive. However, receiving a diploma of recognition from the institution motivated them; they valued it. They considered that the most important merit of their actions was a social merit that they hope will be highlighted when the benefits for the affected patients are achieved.

> *“Yes, they gave me a diploma of donation or participation in this procedure. I did like it, it motivated me, it always generates an internal joy to have some kind of recognition from someone. Actually, I did not expect recognition, but it was good, I liked it”* (JL, man).
>
> *“It is no merit to donate blood, it is no academic merit, it is no merit of life, maybe it is a social and health merit, for helping others, but no more than that. And that doesn’t need papers to be documented”* (MR, man).
>
> *“I tell you something that happened to me, that many people told me “they are not paying you”, “how much have they given you”, “how much has the Hospital paid you” and I tell them nothing, because it is my will to help people, I am not waiting for them to give me something in return for what I do, I hope to be able to help and that my neighbour can recover. The truth is that I don’t agree that there should be recognition, even though I have been given recognition with a diploma, but if you do good, it is to help others and not to expect anything in return”* (DA, man).

### Fears

The main fears mentioned regarding the convalescent plasma donation process were the possibility of fainting due to the effect of blood collection and is located in overcrowded places that are infectious sources of infection during the procedure. However, the participants stated that they found clean, spacious spaces and were accompanied by the health personnel.

> *“I was quite surprised that where they took me out was a spotless, nice place, and it looked good, so I was not afraid. Besides, I told the gentleman to have compassion for me, I am afraid of needles, and the gentleman did have compassion; the gentleman who took my blood was very nice, why not. So when I went to the Rebagliati, I felt confident that they would take me to a specific place to have my blood drawn. I know what hospitals are like in terms of health personnel, I know what they are like inside, how ugly they are; so when they took me to this area that was just for me, I was amazed at how well I was treated and the lady who guided me and everything, she was a good person”* (VR, woman).
>
> *“*… *during the whole procedure, I had the same head of the clinical trial, whom I think is the head of the haematology area, next to me, explaining everything to me, there was a technician attentive to everything. There was no problem at all during the blood donation, or anything else”* (JM, man).
>
> *“At the beginning, yes I was afraid, because I was going to a high-risk place, completely with treatment of infected people. So before arriving, the place itself, what it was like, what it would be like, I was a little afraid of the exposure. But once I got to the place, I had the confidence, because of the staff, the equipment, the environment”* (JL, man).

#### Experiences of convalescent plasma donation

The participants mentioned the positive experiences of the donation process:

1. The punctuality of the medical attention
2. The accompaniment of the health personnel
3. The spacious places for the different activities required for the donation procedure

On the other hand, the most outstanding negative experiences were the problems in relating with some nurses. Second, the length of time from enrolment to blood donation. It is a process that they considered to belong—approximately one month’s duration, which includes registration, testing, and test results. Third, the lack of reserved places for donors. They enter the hospital and wait together with other patients in the general care area until the study’s person comes. However, they considered that given their condition as former COVID patients, this is risky because they represent possible spaces for contagiousness. Fourth, admission to the hospital had an initial negative impact on some participants. The security personnel were not aware of the donation campaign and could not give them a clear orientation on where to go. This misinformation generated some distrust and confusion among participants. Finally, participants mentioned that they are not aware of blood donation campaigns in general that are massively disseminated.

> *“From my blood donation at Rebagliati, the initial experience was not good. Because I arrived, first of all, the security guard did not want to let me in, no matter how much I told him that I came to donate my plasma, that I had already been contacted and everything else. And he told me that he didn’t know anything about COVID plasma donors and that if I had COVID I couldn’t enter, that I should go to the Emergency Room, and I tried to explain to him but he didn’t understand me. So I contacted the lady I contacted and said “look, I’m outside, they won’t let me in”, so she had to go over and get them to let me in”* (ST, female).
>
> *“It seemed very slow to me, a very slow process because I thought that you signed up and I knew that they were going to do a previous test to see your level of defence or immunity, but it seemed slow to me, I think that when someone wants to help they want to do it now, so me. Ah, they told me that something had not arrived, some additives that were needed just the level of defenses or antibodies that I had generated, that is why the process was delayed too much, that is why I say that if they are going to do a campaign of this type, the important thing is the time, that they do it quickly so that other people do not get discouraged”* (MM, man).

### Expectations and recommendations

The donors emphasized that “despite being a public hospital,” they generally received good attention and support throughout the donation process, from enrolment to plasma collection. However, among the main expectations and suggested recommendations, they highlighted the need for greater dissemination of blood donation campaigns in general, both internally (hospital) and externally (community). For example, the promotion of the convalescent plasma donation campaign, from the perception of the participants, did not acquire the expected relevance and amplification. In addition, they suggested that information about the donation process should include messages where safety and detailed procedures are included.

> *“It would be good that when they make the invitations, it would not only be what is needed but that they describe how it is going to be done and what the place is like, that would give people more confidence. We have this equipment, the procedure is extremely sterile, the doctors are protected, things like that. But, because the invitation is cold, you have to donate, and you have to do this, ah ya, but all those doubts are not absolved until you arrive”* (JL, man).
>
> *“It would also be very important to raise awareness among all the people who are related to blood donation because it is very important that the people, from the door, who welcome you to the institution, are related to or know what the people who are going to donate blood are going to do. Because the first time I went to Rebagliati, the procedure was lousy, and I was one step away from leaving, but I remembered and understood that this is how it is often treated”* (ST, woman).
>
> *“My expectations, in relation to donation I think it should be a little more intensive. More frequent campaigns, education in schools I think that would be the best way to look for a better future is with young people and the younger the better, maybe they can’t donate, but they know that at some point in their life they have to do it, it is not unsafe, that it is not a problem, it is a donation and they can help many people, right?”* (CS, male).

On the other hand, another expectation and recommendation focused on hospital infrastructures. Friendly spaces that provide confidence to donors.

> *“*…*I think it should improve especially the sanitation of donation, I have donated in different hospitals and it is always a bit gloomy, a bit like that. Well, when I visit clinic there is an abysmal difference between one place and another. Private clinics, then, definitely the health system has collapsed in the country and I imagine or hope that in the coming years it will be better, if a person is going to make a donation or is going to be part of a process there should be some minimum conditions that fill their eyes so that more people are encouraged, right, a bigger place, a nicer place”* (MM, man).

## Discussion

Although specifically focused on convalescent plasma donation, the study could also reflect part of the reality in terms of blood donation campaigns and the participation of citizens as potential donors. Unfortunately, Perú is one of the countries with the lowest levels of voluntary blood donation in Latin America, with 13.5% in 2019 [13]. According to the Pan American Health Organization, nine out of ten donations are in the country by replacement, made by people seeking to benefit someone in particular (family member or acquaintance) [13]. Therefore, this donation helps specific recipients, leaving the population with little or no social support networks, in situations of vulnerability and with fewer opportunities for recovery. Reversing these figures is fundamental, and the need is even more alarming in cases such as the current global health crisis, where the collapse of health services and the still uncertainty about the development of the pandemic require greater citizen participation and collaboration promote and contribute to research. That said, voluntary blood donation is not only a generous act but also involves altruism, citizenship, and national identity [14].

It should be noted that, given the current global health crisis, donating blood represents a situation of increased risk. In convalescent plasma donors, the problem may be even more complex since they are people who have recently suffered from the disease. Thus, although awareness of the need to overcome the current pandemic may be present in the general population, the suffering and experiences of a patient will not be the same as those of a regular blood donor.

The study’s findings were that the participants were all highly educated and primarily male (9 donors) versus two females. This was due to the selection criteria of the clinical trial, which excluded all women who had had a previous pregnancy (effective or miscarriage). Therefore, we cannot assess the parity of male/female participation in the study. However, other studies in developing countries show that the sociodemographic profile of donors are predominantly male and concentrated in the age group between 30 and 50 years, middle class, upper-middle-class, educated [15–17]. Therefore, it would be necessary to investigate the reasons why the participation of women is less representative. Far from assuming that it falls into a reductionist aspect of wills, it would be interesting to examine and discuss those barriers found by this population group for their participation. Authors coincide in having found that the requirements for donation are related to body weight and size, previous pregnancy, and haemoglobin levels. These aspects place women at a disadvantage when participating in blood donation campaigns [18,19]. In addition, there is a belief that, due to menstruation and anaemia, women are more susceptible to bleeding. This situation negatively affects their experience as donors even when they show greater intention to participate than men [19].

In light of the above, it is essential to consider blood donation as a heterogeneous process that should be built around the donor, considering their sociodemographic variables. Furthermore, recruitment campaigns should include strategies that show effectiveness in reducing adverse reactions and improving the quality of the blood donation experience, which will positively affect the probability of donor return [20,21].

The participants’ educational level may reflect the greater access to information, greater possibilities of transportation, and time to donate [22]. The health crisis caused by the current pandemic impacts people’s physical health and has also deprived part of the population of their jobs, forcing economic and family restructuring. Thus, the fact that most donors have higher levels of education and income makes sense. Another critical point to consider in this regard is that donors with higher levels of education, having experienced the disease, may perceive a lower risk of infection and have greater motivation to donate.

The primary motivations of convalescent plasma donors were, contributed to local research, confidence, and hope for favourable treatments to combat the pandemic and support other patients. These findings are similar to those of other studies, highlighting altruism as the main reason for voluntary donors [21,23,24]. However, it is crucial to discuss whether an altruistic or completely selfless motivation is sustainable. Conjunctural factors, socioeconomic realities, and cultural aspects may converge so that different incentives must support motivations. For example, in some developing countries, incentives may include periodic health check-ups, public recognition, and monetary incentives [15,24,25].

The COVID-19 pandemic has brought to the forefront discussions about the social position of the infected person, the effects on their relationship with the community, and the various constraints they face. As confirmed by various research studies, the lack of knowledge and stigma surrounding COVID-19 forces them to hide their symptoms, avoid seeking medical attention, and getting tested until they are seriously ill. In addition, they do not cooperate in efforts to investigate contacts [26–28]. Stigma is largely fuelled by misinformation rampant in social networks and media, potentially affecting intentions to donate convalescent plasma. Studies have also shown that delays in care for COVID-19 are also due to poor skills to recognise severity symptoms in patients, self-medication (ivermectin), and perception of poor medical competence and unsafe and inadequate quality services [29,30].

The convalescent plasma donors who participated in the study stated that they were not frequent visitors to social security hospitals. However, it is intriguing the ideas of some of them concerning the infrastructures and services offered in the hospital. Prevalent among this population is the expectation that precariousness is synonymous with public health service. This also helps to understand why the possibility of new SARS-CoV-2 infection in the hospital facilities became the main fear among the participants. For this reason, the same participants noted that the attention of health personnel and safety measures were appropriate and suggested greater dissemination of these aspects in donation campaigns. This finding invites interventions by EsSalud to rescue the cleanliness of its facilities for donation campaigns and achieve a positive impact on the participation of the population.

Several studies have shown a drop-in blood donation during the current health crisis [31,32]. In this sense, the present study provides some light on how to increase blood donation in general by promoting and highlighting the safety aspects of hospital institutions. Friendly spaces, safety, security, protection, and more information are necessary components to safeguard patients and blood donors [15,20,21]. Convalescent plasma donors, being former COVID-19 patients, are vulnerable physically and, as noted above, probably socially. Social security should continue to work in this effort to guarantee their care, valuing and recognizing, in addition, their civic contribution to improving the health of others and to the relentless struggle to combat the current health crisis.

### Limitations

The study was conducted with donors who were able to complete the donation process. Therefore, we did not collect testimonies of those who initiated the enrolment and who, for various reasons, either directly related to the requirements of the study or personal nature, did not complete the procedure. Likewise, health personnel involved in convalescent plasma donation were not interviewed.

## Conclusions

The experiences of the convalescent plasma donors in the present study were positive, highlighting the punctuality of medical attention, the accompaniment of health personnel throughout the donation process, and the spacious places for the various activities involved in the donation procedure. However, there are still aspects to improve in terms of processes and infrastructure to ensure successful blood donation campaigns in the future. The time from enrolment to convalescent plasma donation, the relationship with some nurses, the lack of dissemination of the convalescent plasma donation campaign, and the lack of waiting spaces reserved exclusively for former COVID patients were the main observations provided by the participants.

The two main reasons driving the intention to donate convalescent plasma were: i) support for research and ii) support for COVID-19 victims. The role of citizens in the face of health crises is fundamental. Therefore, working on the sustainability of their participation through a comprehensive look at the motivations and barriers of donors, both at the individual and institutional level, should be a continuous work that can guarantee an increase in the population’s participation.

Perú is one of the countries with the lowest rates of voluntary blood donation. Given the current global health crisis, this scenario could be even more complex given the high risks of contagion in various hospital establishments. For this reason, collecting the narratives of convalescent plasma donors has allowed us to explore their experiences both as former COVID patients and as donors and to provide tools and inputs to ensure the safety and confidence of donors in order to amplify citizen participation safely and responsibly.

## Data Availability

All data produced in the present study are available upon reasonable request to the authors.

## References

1. Buzai GD. De Wuhan a Luján. Evolución espacial del COVID-19. 2020 Apr 17 [cited 2020 Nov 19]; Available from: http://ri.unlu.edu.ar/xmlui/handle/rediunlu/683

2. Research C for BE and. Recommendations for Investigational COVID-19 Convalescent Plasma. FDA [Internet]. 2021 Dec 2 [cited 2021 Jul 4]; Available from: https://www.fda.gov/vaccines-blood-biologics/investigational-new-drug-applications-inds-cber-regulated-products/recommendations-investigational-covid-19-convalescent-plasma

3. ¿Es el plasma de las personas que se han recuperado de covid-19 un tratamiento efectivo para las personas con covid-19? [Internet]. [cited 2021 Jun 1]. Available from: /es/CD013600/HAEMATOL_es-el-plasma-de-las-personas-que-se-han-recuperado-de-covid-19-un-tratamiento-efectivo-para-las

4. Piechotta V, Iannizzi C, Chai KL, Valk SJ, Kimber C, Dorando E, et al. Convalescent plasma or hyperimmune immunoglobulin for people with COVID-19: a living systematic review. Cochrane Database of Systematic Reviews [Internet]. 2021 [cited 2021 Jun 1];(5). Available from: https://www.cochranelibrary.com/es/cdsr/doi/10.1002/14651858.CD013600.pub4/full/es

5. International Forum on the Collection and Use of COVID-19 Convalescent Plasma: Protocols, Challenges and Lessons Learned: Summary - Al-Riyami - - Vox Sanguinis - Wiley Online Library [Internet]. [cited 2021 Jun 4]. Available from: https://onlinelibrary.wiley.com/doi/full/10.1111/vox.13113

6. Joyner MJ, Carter RE, Senefeld JW, Klassen SA, Mills JR, Johnson PW, et al. Convalescent Plasma Antibody Levels and the Risk of Death from Covid-19. New England Journal of Medicine. 2021 Mar 18;384(11):1015–27.

7. Libster R, Pérez Marc G, Wappner D, Coviello S, Bianchi A, Braem V, et al. Early High-Titer Plasma Therapy to Prevent Severe Covid-19 in Older Adults. N Engl J Med. 2021 Feb 18;384(7):610–8.

8. Klassen SA, Senefeld JW, Johnson PW, Carter RE, Wiggins CC, Shoham S, et al. The Effect of Convalescent Plasma Therapy on Mortality Among Patients With COVID-19: Systematic Review and Meta-analysis. Mayo Clin Proc. 2021 May;96(5):1262–75.

9. Fisher DL, Alin P, Malnick S. The Evidence for High-Titer Convalescent Plasma in SARS-CoV-2. SN Compr Clin Med. 2021 Mar 1;3(3):790–2.

10. ICTRP Search Portal [Internet]. [cited 2021 Jun 2]. Available from: https://ictrptest.azurewebsites.net/Trial2.aspx?TrialID=PER-013-20

11. OMS | El Perú [Internet]. WHO. World Health Organization; [cited 2021 Jun 2]. Available from: https://www.who.int/workforcealliance/countries/per/es/

12. García Gutiérrez M, Sáenz de Tejada E, Cruz JR. Estudio de factores socioculturales relacionados con la donación voluntaria de sangre en las Américas. Rev Panam Salud Publica. 2003 Mar;13:85–90.

13. SUMINISTRO DE SANGRE PARA TRANSFUSIONES EN LOS PAÍSES DE LATINOAMÉRICA Y DEL CARIBE 2014y2015 [Internet]. [cited 2021 Apr 29]. Available from: https://iris.paho.org/bitstream/handle/10665.2/34082/9789275319581-spa.pdf?sequence=1&isAllowed=y

14. Arias Quispe S, Moscoso Porras M, Matzumura Kasano J, Gutiérrez Crespo H, Pesantes MA. Experiencias y percepciones de los donantes de sangre sobre la donación en un hospital público de Perú. Horizonte Médico (Lima). 2018 Jul;18(3):30–6.

15. Ferguson E, Hill A, Lam M, Reynolds C, Davison K, Lawrence C, et al. A typology of blood donor motivations. Transfusion. 2020;60(9):2010–20.

16. Cimaroli K, Páez A, Bruce Newbold K, Heddle NM. Individual and contextual determinants of blood donation frequency with a focus on clinic accessibility: A case study of Toronto, Canada. Health & Place. 2012 Mar 1;18(2):424–33.

17. Wittock N, Hustinx L, Bracke P, Buffel V. Who donates? Cross-country and periodical variation in blood donor demographics in Europe between 1994 and 2014. Transfusion. 2017;57(11):2619–28.

18. Madrona DP, Herrera MDF, Jiménez DP, Giraldo SG, Campos RR. Women as whole blood donors: offers, donations and deferrals in the province of Huelva, south-western Spain. Blood Transfus. 2014 Jan;12(Suppl 1):s11–20.

19. Bani M, Giussani B. Gender differences in giving blood: a review of the literature. Blood Transfus. 2010 Oct;8(4):278–87.

20. Stephanou AT, Moreira MC. Blood Donors’ Perception of Incentive Campaigns. Paidéia (Ribeirão Preto). 2019;29:e2927.

21. Wevers A, Wigboldus DHJ, de Kort WLAM, van Baaren R, Veldhuizen IJT. Characteristics of donors who do or do not return to give blood and barriers to their return. Blood Transfus. 2014 Jan;12(Suppl 1):s37–43.

22. Javadzadeh Shahshahani H, Yavari MT, Attar M, Ahmadiyeh MH. Knowledge, attitude and practice study about blood donation in the urban population of Yazd, Iran, 2004. Transfus Med. 2006 Dec;16(6):403–9.

23. Asamoah-Akuoko L, Hassall OW, Bates I, Ullum H. Blood donors’ perceptions, motivators and deterrents in Sub-Saharan Africa – a scoping review of evidence. nytgtttftuobbon. 2017;177(6):864–77.

24. Livitz IE, France CR, France JL, Fox KR, Ankawi B, Slepian PM, et al. An automated motivational interview promotes donation intention and self-efficacy among experienced whole blood donors - Livitz - 2019 - Transfusion - Wiley Online Library. [cited 2021 Apr 30]; Available from: https://onlinelibrary.wiley.com/doi/abs/10.1111/trf.15402

25. Mohammed S, Essel HB. Motivational factors for blood donation, potential barriers, and knowledge about blood donation in first-time and repeat blood donors. BMC Hematol. 2018 Dec 20;18:36.

26. Sotgiu G, Dobler CC. Social stigma in the time of coronavirus disease 2019. Eur Respir J. 2020 Aug;56(2):2002461.

27. Abdelhafiz AS, Alorabi M. Social Stigma: The Hidden Threat of COVID-19. Front Public Health [Internet]. 2020 Aug 28 [cited 2021 Apr 30];8. Available from: https://www.ncbi.nlm.nih.gov/pmc/articles/PMC7484807/

28. Villa S, Jaramillo E, Mangioni D, Bandera A, Gori A, Raviglione MC. Stigma at the time of the COVID-19 pandemic. Clinical Microbiology and Infection. 2020 Nov 1;26(11):1450–2.

29. Pajuelo MJ, Anticona Huaynate C, Correa M, Mayta Malpartida H, Ramal Asayag C, Seminario JR, et al. Delays in seeking and receiving health care services for pneumonia in children under five in the Peruvian Amazon: a mixed-methods study on caregivers’ perceptions. BMC Health Serv Res. 2018 Mar 1;18(1):149.

30. Ypanaqué-Luyo P, Martins M. [Utilization of outpatient health services in the Peruvian population]. Rev Peru Med Exp Salud Publica. 2015 Sep;32(3):464–70.

31. García-Erce JA, Romón-Alonso Í, Jericó C, Domingo-Morera JM, Arroyo-Rodríguez JL, Sola-Lapeña C, et al. Blood Donations and Transfusions during the COVID-19 Pandemic in Spain: Impact According to Autonomous Communities and Hospitals. Int J Environ Res Public Health. 2021 Mar 27;18(7).

32. Al Mahmasani L, Hodroj MH, Finianos A, Taher A. COVID-19 pandemic and transfusion medicine: the worldwide challenge and its implications. Ann Hematol. 2021 Feb 1;1–8.

